# More Than Just Arm Movement: Finger-Worn Accelerometers Provide a Valid and Sensitive Alternative to Wrist-Worn Accelerometers for Measuring Real-World Upper-Limb Performance in Stroke Survivors

**DOI:** 10.64898/2026.08.13.26360286

**Authors:** Ramita Dhamrongsirivadh, Benito Lorenzo Pugliese, Virginia Civeriati, Kathy Piela, Eric Fabara, Gloria Vergara-Diaz, Qing Mei Wang, Paolo Bonato, Sunghoon Ivan Lee

## Abstract

**Objective:** To investigate the clinical validity of finger-worn accelerometers for providing a comprehensive assessment of upper-limb motor performance in stroke survivors in real-world environments, compared to wrist-worn accelerometers, and to examine how the clinimetric properties of wearable-based motor performance measures vary with the duration of patient data collection.

**Design:** Cross-sectional observational design.

**Setting:** Research laboratory and free-living environments.

**Participants:** Twenty-seven stroke survivors aged 18-80 years with ischemic or hemorrhagic stroke at least six months prior to enrollment and mild-to-moderate upper-limb impairment without severe range-of-motion restrictions were enrolled. Three participants were ineligible and four withdrew, resulting in a final cohort of 20 participants (N = 20).

**Interventions:** Not applicable.

**Main Outcome Measures:** Wearable-based motor performance measures derived from fine-hand movements, gross-arm movements, and the combination of fine-hand and gross-arm movements captured by finger-worn and wrist-worn accelerometers in naturalistic settings for 6.4 *±* 1.8 days.

**Results:** Wearable-based motor performance measures from fine-hand movements demonstrated the strongest convergent validity, known-group validity, and test-retest reliability, followed by those from combined and gross-arm movements. Convergent validity and test-retest reliability of wearable-based motor performance measures improved with longer monitoring durations, with four days being sufficient to obtain accurate and reliable upper-limb measures.

**Conclusions:** Wearable-based motor performance measures from finger-worn accelerometers provide a more comprehensive assessment of upper-limb motor performance than those from wrist-worn accelerometers, supporting their use for real-world monitoring in stroke survivors. Furthermore, the improvements in clinimetric properties of wearable-based motor performance measures with longer monitoring durations highlight the importance of multi-day monitoring to mitigate day-to-day variability and ensure robust assessment.

---

Stroke affects nearly 800,000 individuals annually in the United States and is a leading cause of long-term disability.^1–2^ Approximately 80% of stroke survivors experience upper-limb motor impairments,^3^ hindering their ability to perform activities of daily living (ADLs). In outpatient stroke rehabilitation, accurate monitoring of real-world upper-limb performance could help guide targeted interventions and assess outcomes in regaining independence in ADLs.^4^

Wrist-worn accelerometers have been widely studied as objective tools for monitoring daily upper-limb use in stroke survivors.^5–6^ However, these devices primarily detect gross-arm movements (upper arm and forearm) and are limited in capturing fine-hand movements (wrist, hand, and finger) associated with ADLs.^7–9^ Consequently, wrist-worn accelerometers may provide an incomplete representation of upper-limb performance, limiting their utility in clinical practice and rehabilitation research.^7,10^

To address these limitations, researchers have investigated alternative upper-limb activity sensing mechanisms. One such approach utilizes magnetometer-embedded wrist-worn devices and magnetic rings to estimate wrist and finger joint angles.^11–12^ However, its feasibility for bilateral monitoring is limited by ferromagnetic interference between sensors, and the need for two sensors per limb increases wearability burden.^13^ As a more promising solution, previous studies have demonstrated that a standalone accelerometer worn on the index finger can overcome these challenges.^9,14–15^ Finger-worn accelerometers detect both gross-arm and fine-hand movements without being affected by environmental noise.^9,14^ Recently, Liu *et al.* showed in a simulated home environment that finger-worn accelerometers may provide stronger convergent validity and sensitivity to impairment levels than wrist-worn devices.^10^ Despite these findings, validation of finger-worn sensors in stroke patients’ real-world environments has not yet been performed.

This study investigates the clinical validity of performance measures derived from finger-worn accelerometers using data collected from stroke survivors in real-world environments. Specifically, we evaluate: 1) convergent validity with standardized clinical assessments of motor capacity and performance, 2) known-group validity in distinguishing impairment levels, and 3) test-retest reliability. Moreover, by collecting sensor data from stroke survivors over multiple days, we examine how monitoring duration influences convergent validity and test-retest reliability. Finally, we conduct a comparative analysis of validity and test-retest reliability against wrist-worn accelerometers.

## Methods

### Participants

Participants were recruited from [Redacted]. Eligibility criteria included: 1) ischemic or hemorrhagic stroke diagnosis ≥6 months prior to enrollment, 2) mild-to-moderate upper-limb impairment (Fugl-Meyer Assessment for Upper Extremity (FMA-UE) >35) without severe range-of-motion restrictions, and 3) age 18-80 years. The FMA-UE threshold excluded individuals with difficulties performing ADLs requiring distal limb function,^16^ as conventional thresholds, such as those proposed by Woodbury *et al.*,^17^ may include a broader impairment spectrum. Exclusion criteria were: inability to lift the affected limb against gravity (<30° flexion and abduction), severe spasticity preventing passive finger movement (Modified Ashworth Scale >3), inability to put on or remove sensors, alone or with caregiver assistance, or cognitive impairment interfering with study procedure comprehension (Mini-Mental State Examination <23). The study protocol was approved by the Institutional Review Board of [Redacted] (IRB [Redacted]).

### Data Collection

Upon laboratory visit, participants underwent clinical motor assessments, including FMA-UE, Wolf Motor Function Test (WMFT), and Motor Activity Log (MAL). The FMA-UE assesses motor impairment in the stroke-affected upper limb.^18^ The WMFT evaluates stroke-affected upper-limb function based on functional ability (FA) and performance time (PT).^19^ The MAL is a semi-structured interview assessing real-world motor performance, capturing amount of use (AOU) and quality of movement (QOM) of the affected limb.^20^

Following the assessments, participants wore custom-designed triaxial accelerometers on both index fingers and wrists (Figure 1) while performing daily activities in naturalistic settings. Finger-worn sensors were sized using a standardized ring-sizing kit, while wrist-worn sensors were secured using wristbands. Sensors continuously recorded acceleration data at a sampling rate of 50 Hz. The intended monitoring period was seven days, but actual data collection varied. Participants removed sensors every night during sleep and avoided wearing them during activities involving significant water exposure.

**Figure 1.**
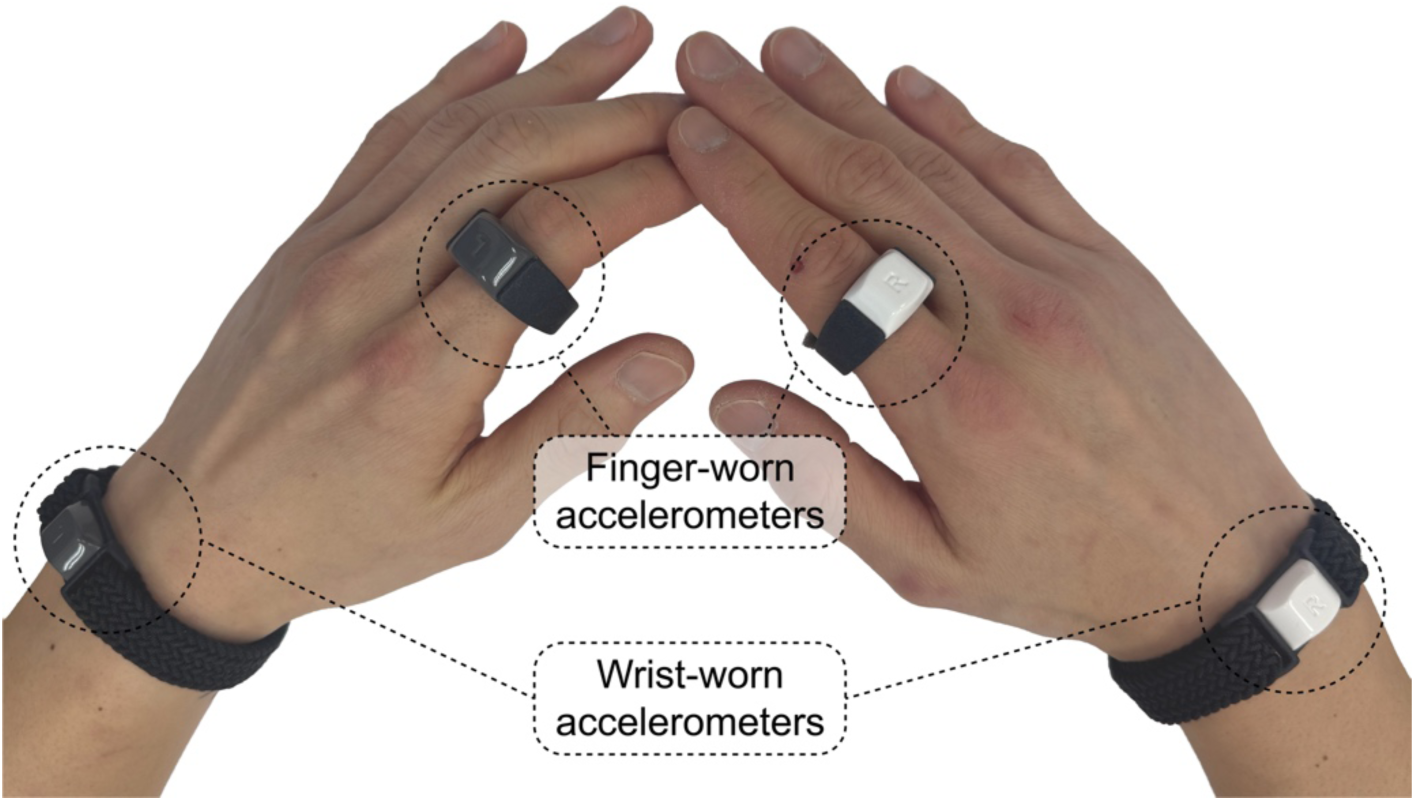
Wrist-worn and finger-worn accelerometers used during data collection.

### Accelerometer Data Pre-Processing

Accelerometer magnitude data from finger-worn and wrist-worn devices were processed to generate three time-series representing: 1) fine-hand movements, 2) gross-arm movements, and 3) the combination of fine-hand and gross-arm movements. Raw acceleration magnitudes were filtered with a sixth-order Butterworth bandpass filter, with cutoff frequencies of 0.1 and 12 Hz, to attenuate gravity and high-frequency noise,^10,21^ and then down-sampled by averaging over one-second epochs.^22^ Only monitoring days with data available from all four sensors were analyzed; within these days, any time segments containing missing or corrupted data from one or more sensors were discarded.

Wrist-worn sensors captured only gross-arm movements; thus, the time-series were labeled 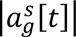 for the stroke-affected limb and 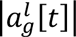 for the contralateral limb. Finger-worn sensors detected a combination of gross-arm and fine-hand movements, resulting in time-series 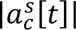 for the stroke-affected limb and 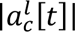 for the contralateral limb. Fine- hand movements were isolated by subtracting wrist-worn sensor data from finger-worn sensor data using the following equation:

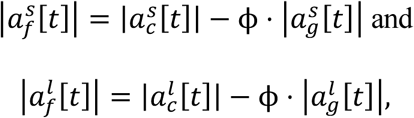

where *t* is in seconds, and *ϕ* is a scaling constant compensating for acceleration differences due to distinct sensor locations. This adjustment ensured isolation of fine-hand movements rather than simply reflecting greater acceleration at the more distal site.

To determine *ϕ*, the forearm was modeled as a pendulum with the elbow as the fulcrum, resulting in identical angular acceleration at both sensor sites. However, linear acceleration is greater at the finger due to its increased distance from the fulcrum. To correct for this, *ϕ* was calculated as the ratio of the distances from the elbow to each sensor: from the elbow to the metacarpophalangeal joint of the middle finger (finger-worn sensor) and from the elbow to the wrist (wrist-worn sensor). These distances were derived from population-based anthropometric data rather than participant-specific measurements to ensure scalability in real-world deployments.^23,24^ Consequently, *ϕ* was set to 36.03 cm / 26.25 cm *≈* 1.372 for female participants and 40.14 cm / 29.03 cm *≈* 1.383 for male participants.

This procedure yielded three pairs of acceleration time-series, representing fine-hand movements 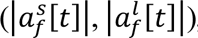, gross-arm movements 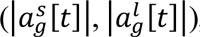, and combined movements 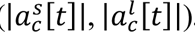.

### Wearable-Based Assessments of Upper-Limb Motor Performance

From each pair of fine-hand, gross-arm, and combined movement time-series, we extracted three broadly used, wear-time-independent measures of upper-limb activity: 1) average intensity of the stroke-affected limb,^25^ 2) percentage duration of active use of the stroke-affected limb,^22^ and 3) use ratio.^26–27^ Average intensity of the stroke-affected limb (*I^s^*) was calculated as the mean acceleration magnitude across all epochs during the monitoring period:

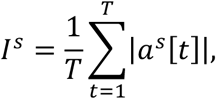

where *T* is the total number of one-second epochs.

Percentage duration of active use (*D^s^*) represents the proportion of epochs during which the stroke-affected limb was active:

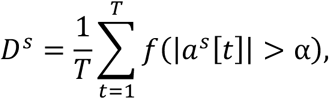

where *f* (*·*) returns 1 if the input condition is met and 0 otherwise, and *α* is an acceleration threshold for identifying active limb use. While studies often use *α* = 0.003328 *g* as a standard threshold,^28–29^ we optimized *α* using a data-driven approach by performing a grid search to maximize the correlation between percentage duration of active use and FMA-UE. FMA-UE was selected as the reference based on its established use for evaluating convergent validity of wearable-based performance measures.^30–31^ To avoid overfitting, threshold selection was performed using leave-one-subject-out cross-validation (LOSOCV). For each iteration, the optimal threshold *α* was determined from the training participants and applied to the held-out participant. Separate thresholds were determined for fine-hand, gross-arm, and combined time-series to account for location-specific acceleration differences.

Use ratio (*R^u^*) quantifies relative active use of the stroke-affected limb compared to the contralateral limb, calculated as the logarithmic ratio of active use durations:

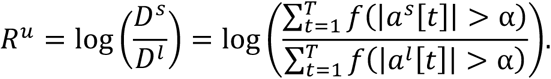

This resulted in nine motor performance measures per participant: *I^s^*, *D^s^*, and *R^u^* from fine-hand, gross-arm, and combined time-series.

### Statistical Analysis

We evaluated three clinimetric properties of wearable-based motor performance measures: 1) convergent validity, 2) known-group validity, and 3) test-retest reliability. Additionally, we examined how monitoring duration influenced convergent validity and test-retest reliability by simulating shorter data collection periods with data available for *k* days.

For each simulated duration *k*, we computed wearable measures and their clinimetric properties. For convergent validity, we selected 100 random combinations of consecutive *k* days from the full dataset. Mean and standard deviation of convergent validity were reported across all combinations. For participants with <*k* days of data, measures were derived from their longest available monitoring period.

### Assessment of Clinimetric Properties

Convergent validity was evaluated using Spearman correlation coefficients between wearable measures and standardized clinical assessments, including FMA-UE, WMFT, and MAL. For known-group validity, participants were categorized into mildly and moderately impaired groups using an FMA-UE cutoff of 47.^17^ Between-group differences were tested using the Mann-Whitney test to determine whether wearable measures could distinguish impairment levels.^32^ Within-group comparisons of measures from the three time-series (fine-hand, gross-arm, and combined) were performed using the Wilcoxon signed-rank test.^33^ Effect sizes were reported using Cliff’s δ.^34–35^

Test-retest reliability was quantified using intraclass correlation coefficients (ICC(2,1)), derived from pairs of wearable measures from consecutive, non-overlapping monitoring periods. For one-day monitoring intervals, test-retest reliability was assessed by comparing measures across two successive days (e.g., day 1 vs. day 2, day 2 vs. day 3). For multi-day intervals (e.g., three days), test-retest reliability was evaluated by comparing aggregate measures from two consecutive periods (e.g., days 1–3 vs. days 4–6).

Convergent validity was evaluated for monitoring durations up to six days. In contrast, test-retest reliability was restricted to intervals up to three days, as calculation required at least two consecutive blocks within the six-day period (e.g., days 1-3 vs. days 4-6).

## Results

Twenty-seven chronic stroke survivors with mild-to-moderate upper-limb impairment were enrolled. Three participants were ineligible and four withdrew, resulting in a final cohort of 20 participants. Participant demographics and clinical assessment scores are summarized in Table 1 (see Supplemental Table S1 for participant-level data). All participants completed at least the intended seven days of data collection. After handling missing or corrupted data, a total of 128 days were included, averaging 6.4 *±* 1.8 days/participant. Of these, 96% of acceleration data were retained, corresponding to 1,158 hours and an average wear time of 9.1 *±* 4.5 hours/day (Supplemental Table S2).

**Table 1.** Participant demographics and standardized clinical assessment scores.

|  | All | Moderate | Mild |
| --- | --- | --- | --- |
|  | (n = 20) | (n = 9) | (n = 11) |
| **Age; Median (IQR)** | 63 (9.75) | 67 (10) | 62 (8) |
| **Sex** | 12 males, 8 females | 6 males, 3 females | 6 males, 5 females |
| **Chronicity; Median (IQR)** | 36.5 (61.75) months | 52 (75) months | 21 (47) months |
| **Dominant Side** | 7 left, 13 right | 3 left, 6 right | 4 left, 7 right |
| **Affected Side** | 9 left, 11 right | 5 left, 4 right | 4 left, 7 right |
| **FMA-UE; Mean ± SD** |  |  |  |
| Upper-Extremity | 27.80 ± 4.75 | 24.44 ± 3.97 | 30.55 ± 3.45 |
| Coordination & Speed | 3.95 ± 1.36 | 3.33 ± 0.71 | 4.45 ± 1.57 |
| Wrist | 7.45 ± 2.06 | 5.78 ± 1.79 | 8.82 ± 0.98 |
| Hand | 11.35 ± 3.27 | 8.78 ± 3.35 | 13.45 ± 0.69 |
| Total | 50.55 ± 8.87 | 42.33 ± 4.18 | 57.27 ± 5.00 |
| **WMFT; Mean ± SD** |  |  |  |
| FAS | 59.06 ± 10.34 | 50.63 ± 8.12 | 65.80 ± 6.09 |
| TIME | 2.94 ± 2.09 | 4.44 ± 2.43 | 1.84 ± 0.79 |
| **MAL; Mean ± SD** |  |  |  |
| AOU | 2.86 ± 1.47 | 1.76 ± 0.95 | 3.86 ± 1.09 |
| QOM | 2.78 ± 1.23 | 1.78 ± 0.90 | 3.69 ± 0.61 |

### Acceleration Thresholds for Identifying Active Limb Use

The optimal acceleration thresholds *α* for identifying active limb use across participants were 0.057 *±* 0.019 g, 0.054 *±* 0.003 g, and 0.03 *±* 0.001 g for combined, gross-arm, and fine-hand movement time-series, respectively (Supplemental Figure S1). The optimal threshold for gross-arm time-series was higher than the standard threshold of 0.003328 g^28,29^ and the 0.033446 g (20.1 activity counts) threshold reported by Pohl *et al.*^36^, implying reduced sensitivity to low-intensity movements. These differences may reflect our longer data collection period, which captured a broader range of real-world activities and therefore required a different threshold.

### Convergent Validity with Standardized Clinical Assessments

Table 2 summarizes Spearman correlations between wearable-based motor performance measures and standardized clinical assessments (see Supplemental Table S3 and Supplemental Figure S2 for wearable-based measure distributions). Measures from fine-hand time-series showed the strongest correlation with FMA-UE, followed by combined and gross-arm time-series (Figure 2). For FMA-UE subcomponents, fine-hand time-series yielded the strongest correlations with the wrist and hand components. In contrast, measures from all time-series showed comparable moderate correlations with the upper extremity component, and weak correlations with the coordination and speed component. For other clinical assessments, fine-hand and combined time-series generally demonstrated comparable strong correlations. The exception was use ratio, where gross-arm time-series showed the strongest correlations with WMFT-TIME and MAL-AOU, though marginally stronger than fine-hand and combined time-series.

**Figure 2.**
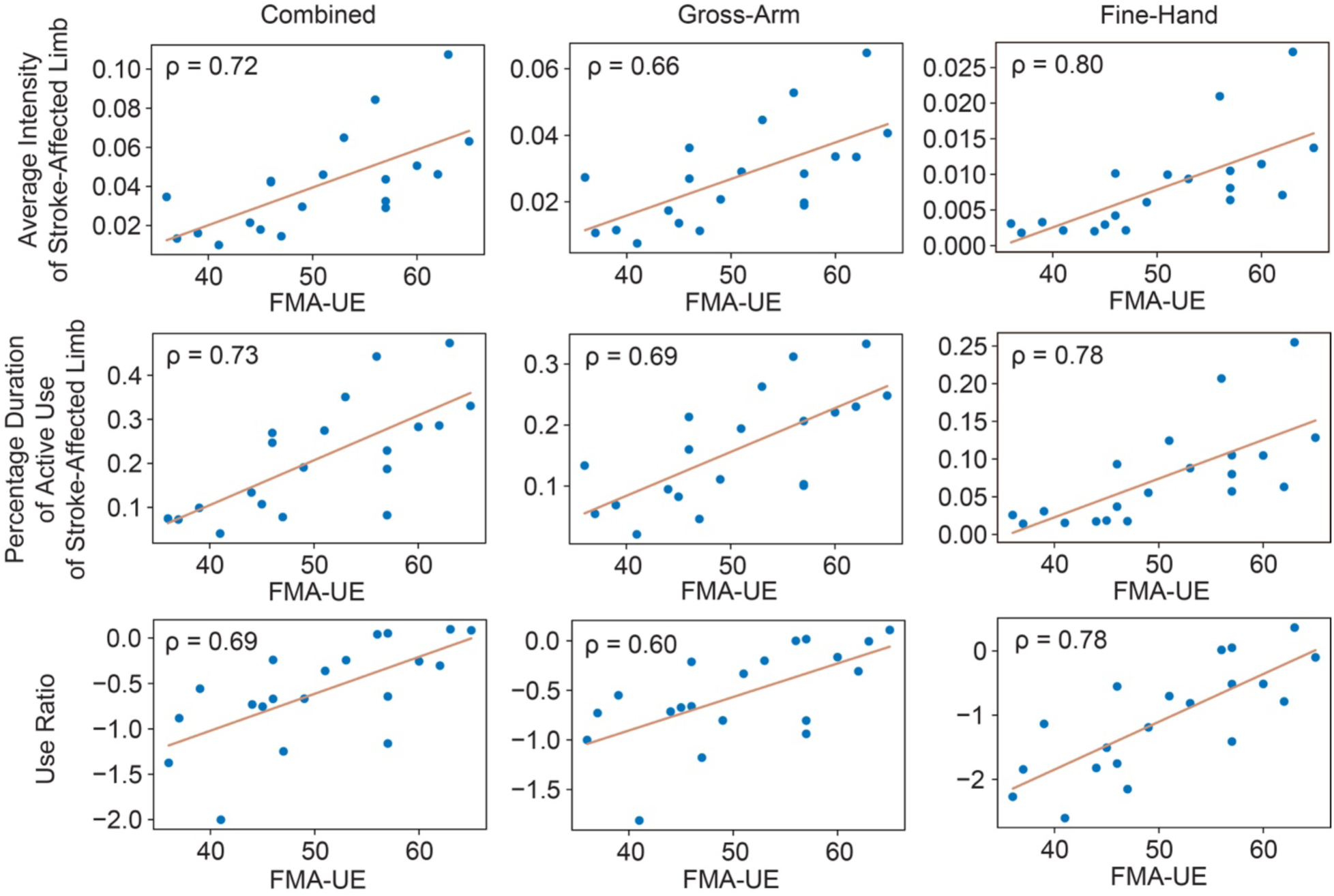
Scatter plots between wearable-based motor performance measures and FMA-UE.

**Table 2.** Spearman correlation coefficients between wearable-based motor performance measures and standardized clinical assessments. Bold formatting indicates the strongest correlation coefficients among the combined, gross-arm, and fine-hand movement time-series. Since shorter task completion times reflect better upper-limb performance,37 negative correlations were found between wearable measures and WMFT-TIME.

| Wearable-Based<br>Motor Performance<br>Measures | Movement<br>Time-Series | FMA-UE |  |  |  |  | WMFT |  | MAL |  |
| --- | --- | --- | --- | --- | --- | --- | --- | --- | --- | --- |
|  |  | Upper<br>Extremity | Coordination<br>& Speed | Wrist | Hand | Total | FAS | TIME | AOU | QOM |
| Average Intensity of<br>Stroke-Affected Limb | Combined | 0.64 | 0.30 | 0.72 | 0.54 | 0.72 | **0.83** | **-0.64** | 0.74 | **0.70** |
|  | Gross-Arm | 0.62 | 0.23 | 0.64 | 0.45 | 0.66 | 0.77 | -0.58 | 0.65 | 0.61 |
|  | Fine-Hand | 0.60 | 0.42 | 0.80 | 0.73 | **0.80** | **0.83** | -0.59 | **0.75** | 0.69 |
| Percentage Duration of<br>Active Use of Stroke-Affected Limb | Combined | 0.59 | 0.40 | 0.72 | 0.55 | 0.73 | **0.85** | **-0.67** | **0.77** | **0.76** |
|  | Gross-Arm | 0.64 | 0.26 | 0.69 | 0.50 | 0.69 | 0.82 | -0.64 | 0.72 | 0.68 |
|  | Fine-Hand | 0.57 | 0.47 | 0.79 | 0.71 | **0.78** | 0.84 | -0.63 | 0.76 | 0.73 |
| Use Ratio | Combined | 0.52 | 0.35 | 0.70 | 0.59 | 0.69 | **0.93** | -0.73 | 0.72 | 0.73 |
|  | Gross-Arm | 0.52 | 0.22 | 0.61 | 0.47 | 0.60 | 0.89 | **-0.75** | **0.78** | 0.70 |
|  | Fine-Hand | 0.61 | 0.37 | 0.79 | 0.72 | **0.78** | 0.90 | -0.68 | 0.76 | **0.75** |

### Known-Group Validity in Distinguishing Impairment Levels

Figure 3 compares wearable-based motor performance measures between moderately and mildly impaired groups. Combined time-series yielded the highest average intensity (Figure 3a) and percentage duration of active use (Figure 3b), followed by gross-arm and fine-hand time-series. This hierarchy supports construct validity of the measures: the combined signal captures the summation of gross-arm and fine-hand activities. All measures significantly differed between impairment groups (*p <* 0.05); however, Cliff’s δ was greatest for fine-hand time-series. For average intensity and use ratio, combined time-series yielded higher Cliff’s δ than gross-arm time-series.

**Figure 3.**
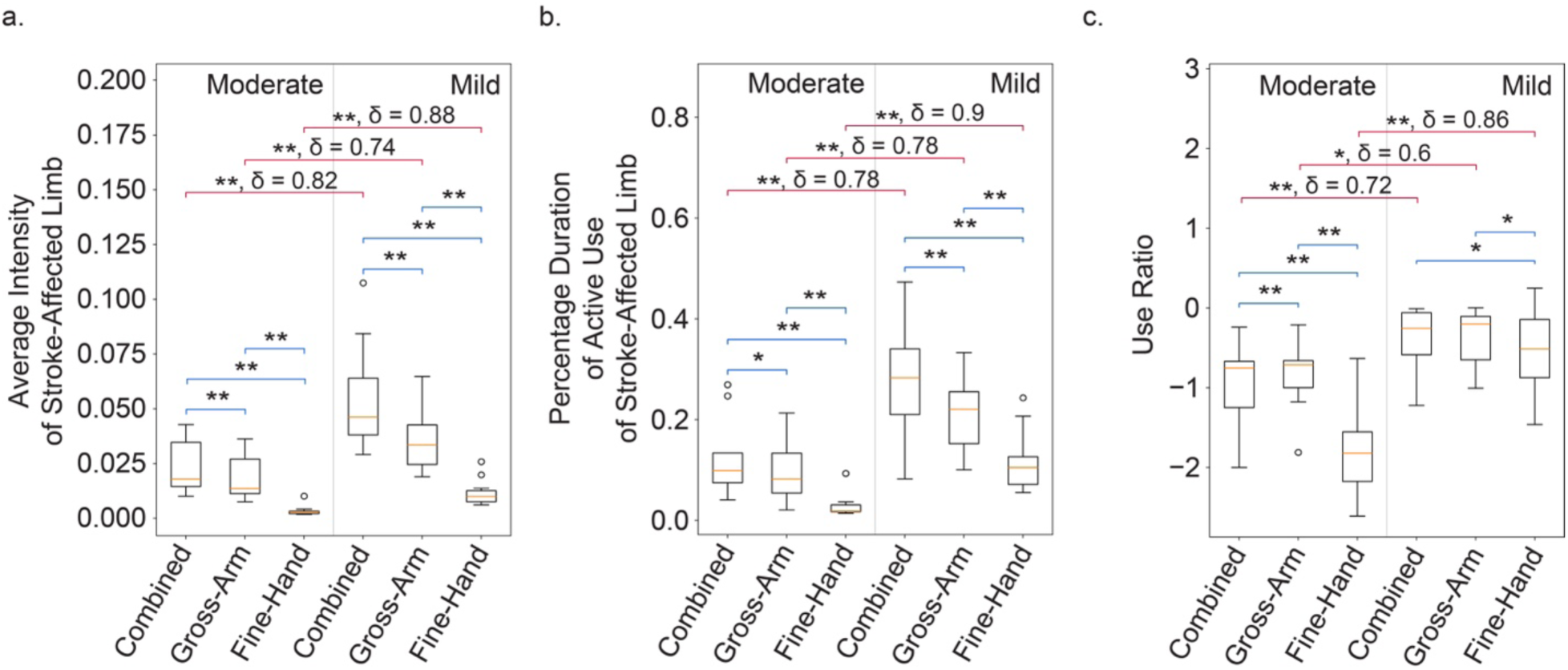
Comparisons of wearable-based motor performance measures from combined, gross-arm, and fine-hand movement time-series between different impairment groups. Blue brackets denote significance from the Wilcoxon signed-rank test, and red brackets indicate significance from the Mann-Whitney U test. A single asterisk (*) represents statistical significance at p < 0.05, while a double asterisk (**) indicates p < 0.01.

Regarding use ratio (Figure 3c), fine-hand time-series exhibited significantly lower values than gross-arm and combined time-series, highlighting more effectively asymmetry in upper-limb use. This likely reflects the well-documented recovery pattern, in which there is a significantly higher prevalence of individuals who regain or retain proximal control (arm and forearm) than distal control (fingers and hand).^38–39^ Consequently, finger-worn sensors revealed functional deficits and asymmetry in the moderately impaired group that were masked when monitoring only gross-arm activity. This is further supported by the effect size analysis, where fine-hand measures demonstrated the largest effects, followed by combined and gross-arm time-series. These results collectively indicate that fine-hand measures provide the greatest sensitivity to motor performance differences across impairment levels.

### Effects of Monitoring Duration on Convergent Validity and Test-Retest Reliability

Figure 4 presents mean and standard deviation of Spearman correlations between wearable-based measures and FMA-UE (top), along with ICC(2,1) values (bottom), across simulated monitoring durations. Both correlation and ICC values increased with longer durations. Correlations plateaued by four days for all measures, whereas ICC values plateaued by two days only for use ratio.

**Figure 4.**
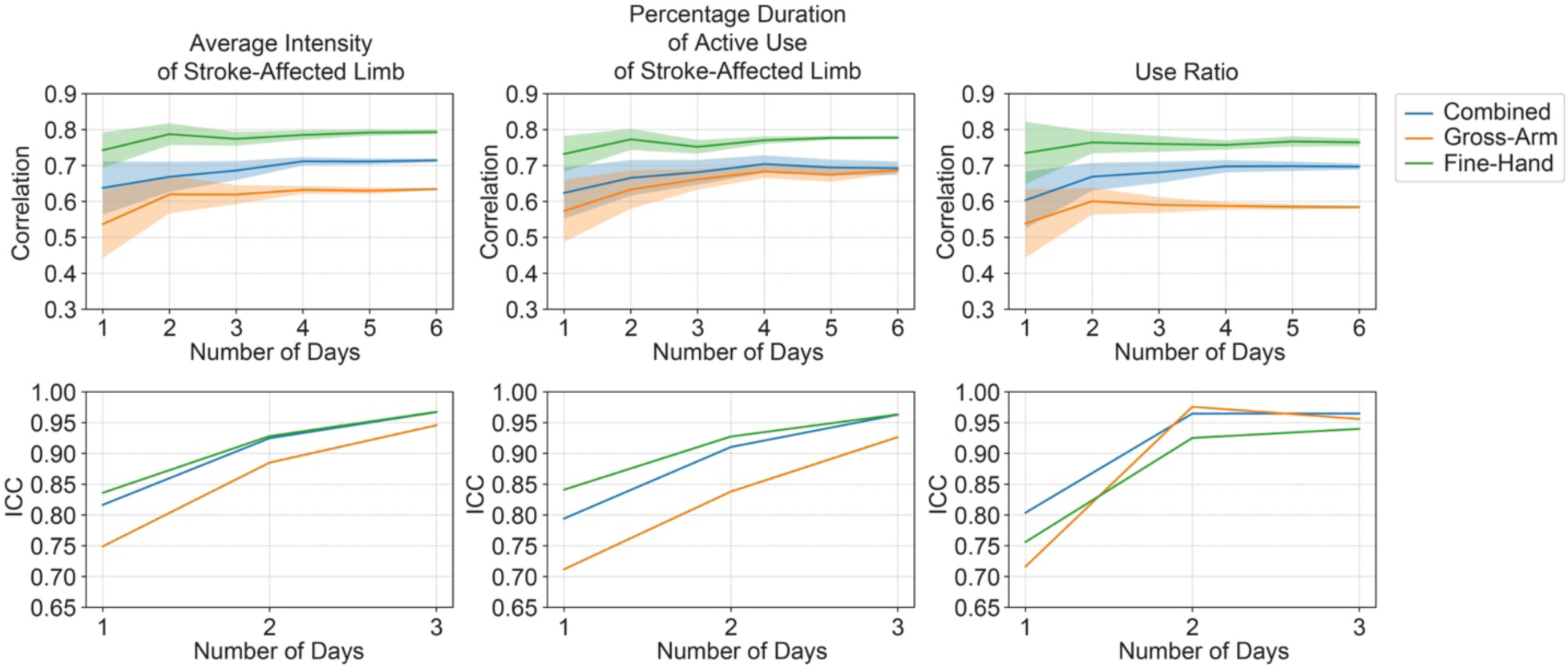
(a) The mean and standard deviation of Spearman correlation coefficients between wearable-based motor performance measures and FMA-UE, and (b) ICC values as a function of monitoring duration.

Across simulated durations, fine-hand measures consistently showed the strongest correlations, followed by combined and gross-arm time-series. A similar pattern was observed for ICC values, except for use ratio. For this metric, combined and gross-arm time-series plateaued earlier (at two days) than fine-hand time-series. Nevertheless, by day three, test-retest reliability of use ratio converged to similar values across all time-series.

Because data were aggregated over monitoring periods regardless of day type (weekdays vs. weekends), we investigated whether wearable measures differed between these periods. No significant differences were found between measures averaged over weekdays versus weekends using paired-samples *t*-tests (Supplemental Figure S3).

### Robustness of Statistical Estimates to Sampling Variability

The reported correlation and ICC values were robust to sampling variability based on jackknife sensitivity analysis (Supplementary Material).

## Discussion

This study investigated the clinical validity of finger-worn accelerometers for assessing upper-limb motor performance in stroke survivors, compared to wrist-worn devices. Measures from fine-hand movement time-series—obtained by analytically subtracting wrist acceleration from finger acceleration—demonstrated the strongest convergent validity, known-group validity, and test-retest reliability, followed by measures from combined (finger acceleration) and gross-arm (wrist acceleration) time-series. Additionally, we examined the effect of monitoring duration on convergent validity and test-retest reliability of wearable measures. The convergent validity and test-retest reliability improved with longer durations, with four days appearing sufficient for accurate, reliable assessment of upper-limb activity.

Convergent validity results (Figure 2 and Table 2) indicate that fine-hand time-series most accurately reflects motor impairments as measured by FMA-UE. Because stroke survivors may not show good recovery of fine-hand function despite improvements in gross-arm function,^18,38^ fine-hand activity reflects greater variability in motor performance among individuals with mild-to-moderate impairment. Consequently, it captures a closer association with wrist and hand impairments, evidenced by stronger correlations with FMA-UE wrist and hand components. This sensitivity may contribute to the increased correlations with total FMA-UE. Moreover, while combined time-series demonstrated weaker yet comparable validity to fine-hand time-series, gross-arm time-series from wrist-worn accelerometers yielded inferior performance to both.

Similarly, fine-hand time-series demonstrated the strongest sensitivity to motor performance differences between moderately and mildly impaired stroke survivors (Figure 3), followed by combined and gross-arm time-series. This finding aligns with results reported by Liu *et al.*,^10^ who validated finger-worn accelerometer data in a laboratory setting. Collectively, these results suggest that finger-worn sensors capture subtle yet meaningful recovery patterns not detected by wrist-worn sensors.

Improvements in convergent validity and test-retest reliability with longer monitoring durations (Figure 4) highlight the necessity of multi-day monitoring to mitigate daily variations and ensure robust assessment. This aligns with established measurement theory^40^ and empirical evidence^41^ that aggregating multiple measurements reduces error and enhances test-retest reliability. While convergent validity plateaued within four days for all measures, test-retest reliability exhibited different patterns: use ratio stabilized by day two, whereas average intensity and active use duration improved through day three. The rapid stabilization of use ratio likely reflects its robustness to physical activity fluctuations.^42^ Nevertheless, the substantially slower improvements for intensity and duration by day three suggest that test-retest reliability likely stabilizes by day four. Furthermore, all measures already achieved excellent test-retest reliability by day three.^43^ Consequently, a four-day monitoring period appears sufficient for valid, reliable assessments, aligning with studies reporting that 3-6 days are needed to reliably capture physical activity in healthy adults using accelerometers.^44–45^

Although fine-hand time-series demonstrated the strongest convergent validity, known-group validity, and test-retest reliability, isolating these movements requires a four-sensor setup (a wrist and finger unit on each limb), which may be burdensome for long-term monitoring. In contrast, combined time-series from finger-worn sensors alone demonstrated performance comparable to fine-hand measures and superior to gross-arm measures from wrist-worn sensors. Therefore, finger-worn sensors may offer a practical balance between clinimetric precision and patient comfort. By providing accurate, continuous real-world monitoring of upper-limb function, these devices could complement conventional clinical assessments and support outpatient rehabilitation.

### Study Limitations

This study has several limitations. First, the small sample size and inclusion of only stroke survivors with mild-to-moderate impairment limit generalizability. For individuals with severe impairment, who typically exhibit minimal fine-hand movement, results may differ; in such cases, wrist-worn sensors may suffice to assess upper-limb motor performance. Furthermore, because of their lower acceleration magnitudes,^46–47^ our optimal acceleration thresholds *α* may not generalize to individuals with severe impairment. Second, isolating fine-hand movements using population-based anthropometric data rather than participant-specific measurements introduces estimation error and noise. Incorporating participant-specific measurements could further improve accuracy. Third, variability in data collection duration and missing or corrupted data may have influenced the results. More consistent and complete data collection would strengthen future analyses. Finally, further research is needed to evaluate the responsiveness of finger-worn accelerometers to longitudinal changes.

## Conclusions

This study investigated the clinical validity of finger-worn accelerometers for comprehensive assessment of upper-limb motor performance in stroke survivors in real-world environments. By capturing both gross-arm and fine-hand movements, finger-worn accelerometers offer stronger convergent validity, known-group validity, and test-retest reliability than wrist-worn accelerometers. Furthermore, our findings indicate that convergent validity and test-retest reliability improve with longer monitoring durations, with four days sufficient to achieve accurate, reliable assessment of upper-limb performance. These results highlight the potential of finger-worn accelerometers for real-world monitoring of upper-limb performance to evaluate rehabilitation outcomes and inform targeted interventions.

## Supporting information

Jackknife Analysis

## Declaration of generative AI and AI-assisted technologies in the manuscript preparation process

During the preparation of this work the authors used ChatGPT and Gemini in order to improve readability and language. After using these tools/services, the authors reviewed and edited the content as needed and take full responsibility for the content of the published article.

## Abbreviations

ADL: Activities of Daily Living
AOU: Amount of Use
FA: Functional Ability
FMA-UE: Fugl-Meyer Assessment for Upper Extremity
ICC: Intraclass Correlation Coefficient
LOSOCV: Leave-One-Subject-Out Cross-Validation
MAL: Motor Activity Log
PT: Performance Time
QOM: Quality of Movement
WMFT: Wolf Motor Function Test

## Data Availability

The data in this study are not publicly available due to privacy and ethical restrictions.

## Supplemental Figure Legends

**Figure S1**. Means and standard deviations of the Spearman correlation coefficients between the percentage duration of active use of the stroke-affected limb and FMA-UE (solid line; left y-axis) and the percentage duration of active use of the stroke-affected limb (dashed line; right y-axis) across participants as a function of the acceleration threshold *α*. The triangle marks indicate the mean optimal values of *α* for each movement time-series across the iterations of LOSOCV.

**Figure S2**. Distributions of the wearable-based motor performance measures.

**Figure S3**. Box plots comparing wearable-based motor performance measures during weekdays and weekends.

**Figure S4**. (a) The mean and standard deviation of Spearman correlation coefficients between wearable-based motor performance measures and FMA-UE, and (b) ICC values as a function of monitoring duration across iterations of jackknife sensitivity analysis.

## Notes

### Competing Interest Statement

The authors have declared no competing interest.

### Author Declarations

IRB of Mass General Brigham gave ethical approval for this work.

### Funding

- NIH: 5R01EB027777
- NSF-NIH Smart and Connected Health Program: R01HD114147

