## Supplementary figures and images for "More Than Just Arm Movement: Finger-Worn Accelerometers Provide a Valid and Sensitive Alternative to Wrist-Worn Accelerometers for Measuring Real-World Upper-Limb Performance in Stroke Survivors"

### Supplemental Figure S1

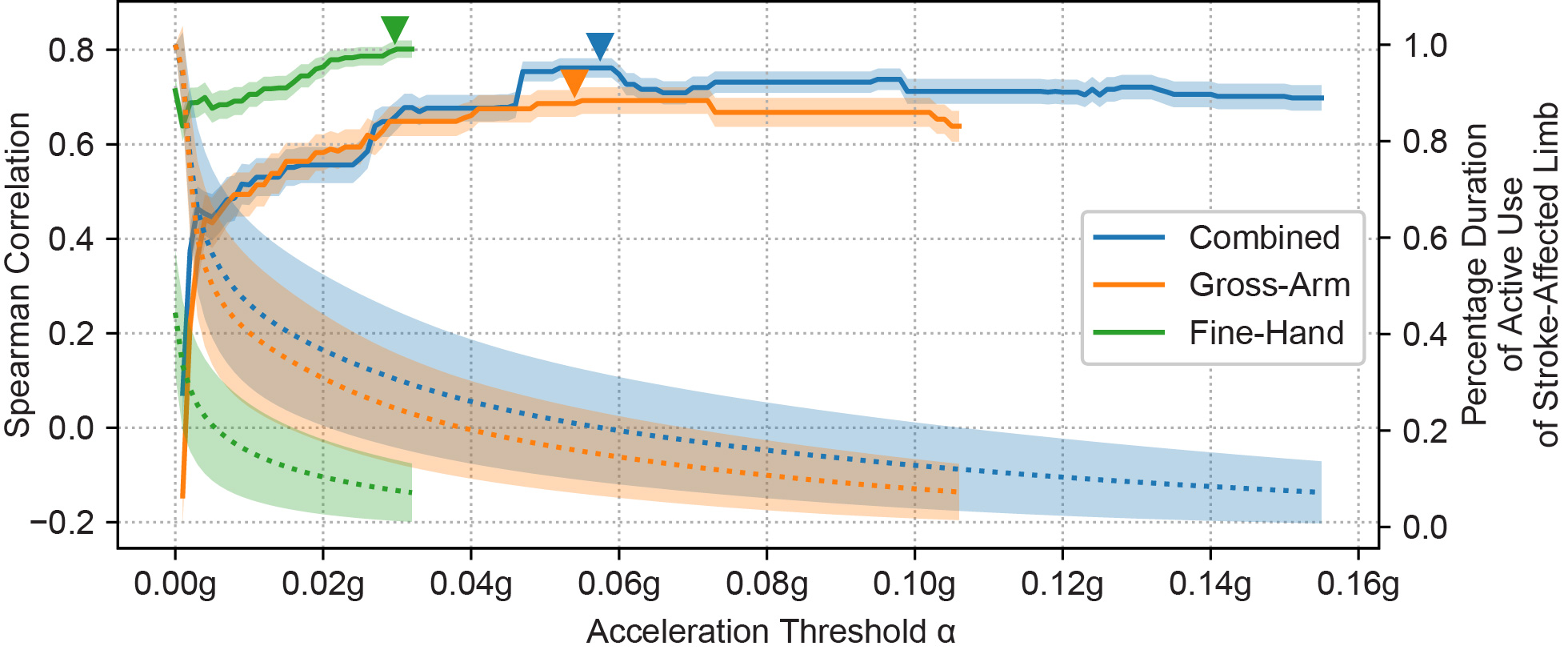

### Supplemental Figure S2

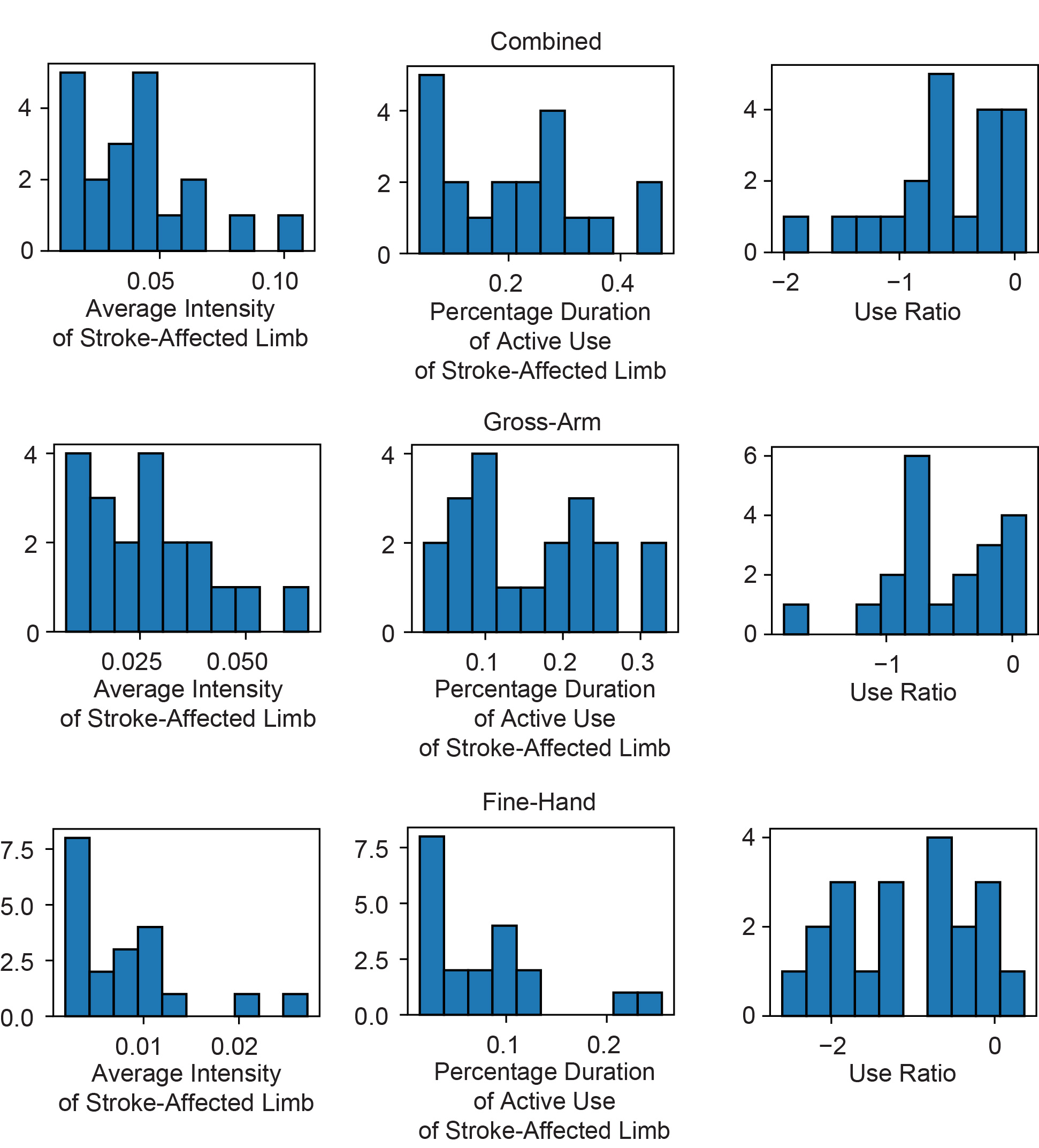

### Supplemental Figure S3

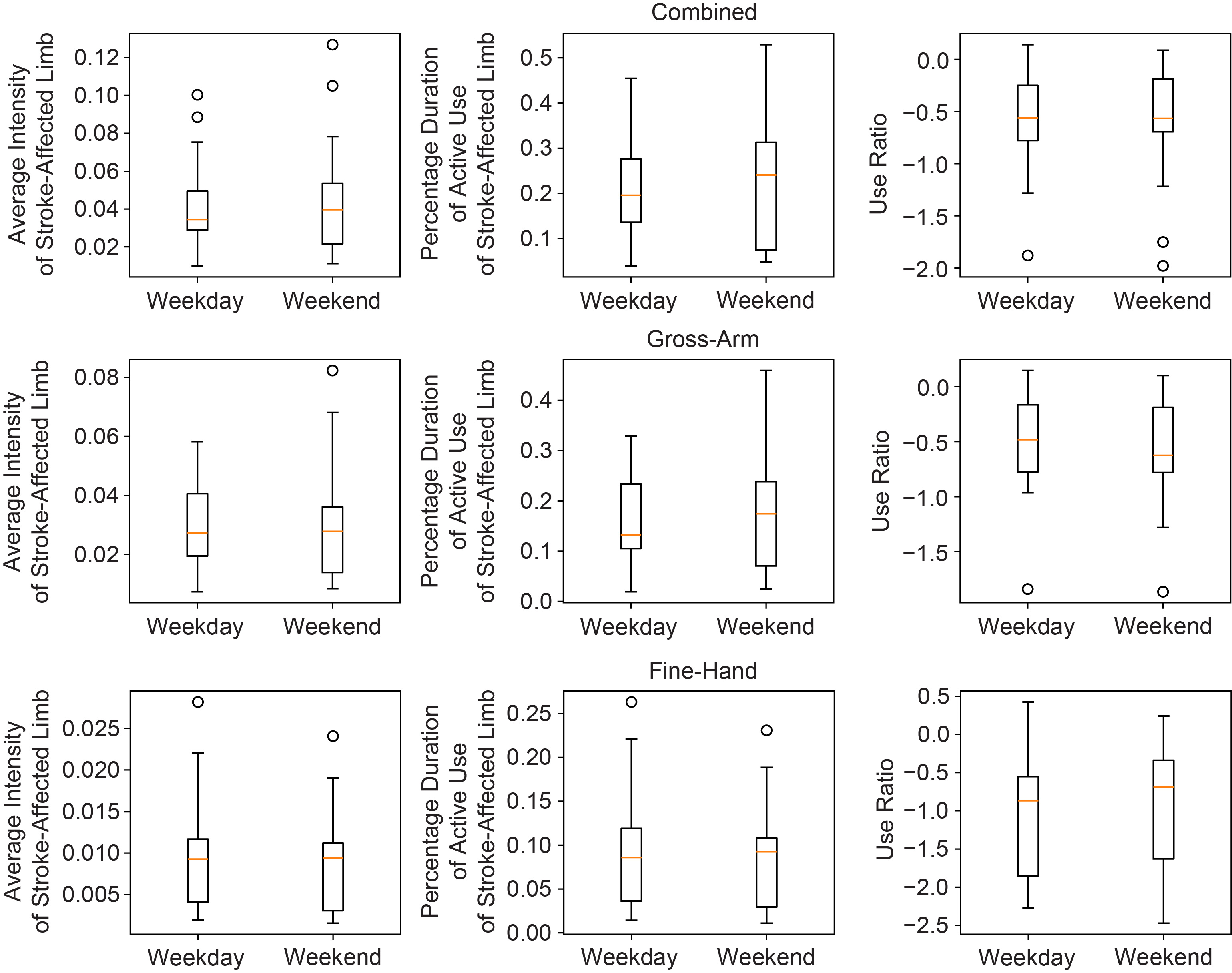

### Supplemental Figure S4

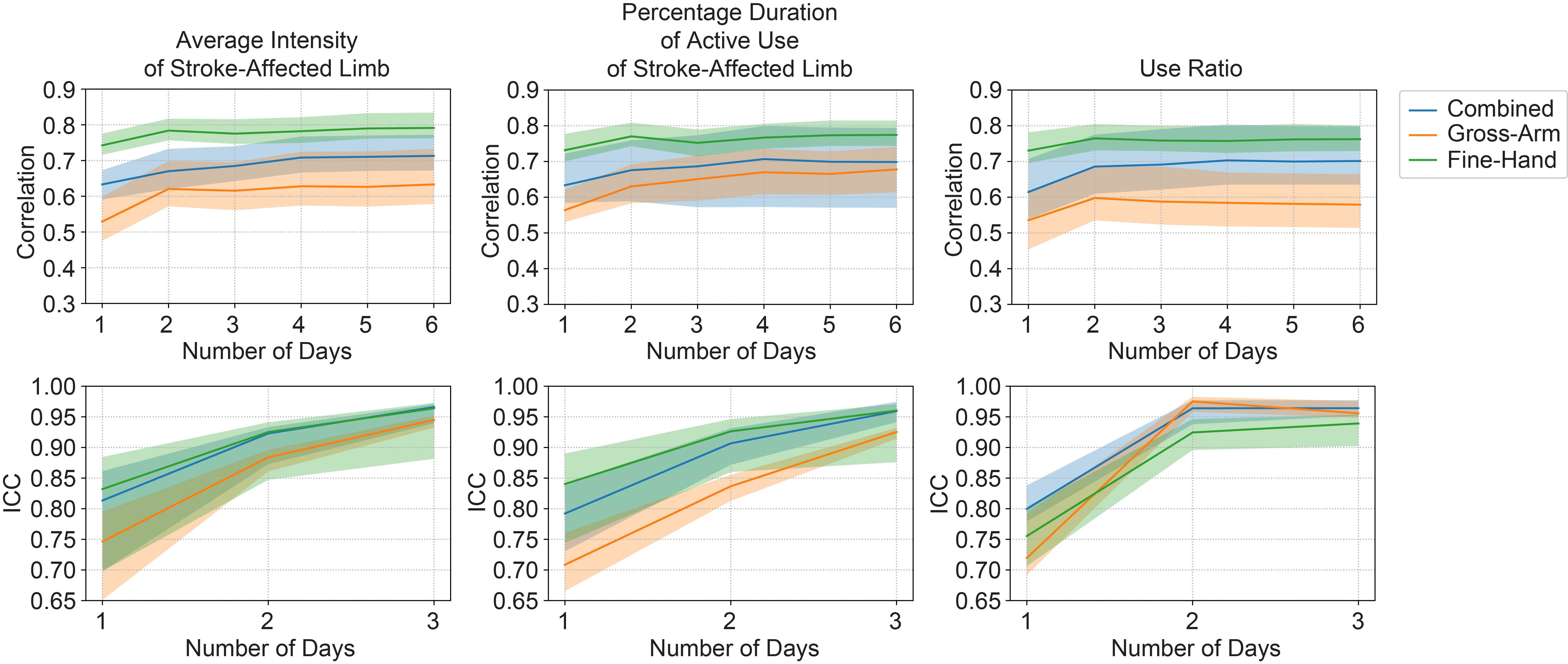
