## Supplemental Table S1 for "More Than Just Arm Movement: Finger-Worn Accelerometers Provide a Valid and Sensitive Alternative to Wrist-Worn Accelerometers for Measuring Real-World Upper-Limb Performance in Stroke Survivors"

Table S1: Individual participant demographics and standardized clinical assessment scores. Missing values for clinical assessments were due to clinician error or participant non-completion.

| **ID** | **Sex** | **Chronicity**  **(Months)** | **Dominant**  **Side** | **Affected**  **Side** | **FMA-UE** | | | | | **WMFT** | | **MAL** | |
| --- | --- | --- | --- | --- | --- | --- | --- | --- | --- | --- | --- | --- | --- |
|  |  |  |  |  | **Upper**  **Extremity** | **Coordination**  **& Speed** | **Wrist** | **Hand** | **Total** | **FAS** | **TIME** | **AOU** | **QOM** |
| **RSA201** | Female | 6 | Right | Left | 35 | 2 | 9 | 14 | 60 |  | 2.91 | 4.76 | 2.48 |
| **RSA202** | Female | 246 | Right | Right | 21 | 4 | 5 | 11 | 41 | 34 | 8.22 | 0.77 | 0.83 |
| **RSA203** | Male | 52 | Left | Left | 29 | 4 | 7 | 5 | 45 | 51 | 2.81 | 1.47 | 2.17 |
| **RSA204** | Female | 49 | Right | Right | 35 | 6 | 10 | 14 | 65 | 72 | 1.12 | 4.77 | 4.43 |
| **RSA205** | Male | 36 | Right | Left | 20 | 3 | 3 | 11 | 37 | 54 | 2.43 | 2.03 | 2.07 |
| **RSA206** | Male | 20 | Left | Right | 33 | 6 | 10 | 14 | 63 | 75 | 1.84 | 3.47 | 3.47 |
| **RSA207** | Male | 99 | Right | Right | 19 | 3 | 4 | 13 | 39 | 57 | 2.51 | 2.3 | 2.13 |
| **RSA208** | Male | 21 | Right | Right | 29 | 3 | 8 | 13 | 53 | 65 | 1.47 | 3.97 | 3.83 |
| **RSA209** | Male | 76 | Right | Left | 26 | 4 | 7 | 12 | 49 | 60 | 1.97 | 1.21 | 3.04 |
| **RSA210** | Female | 359 | Left | Right | 30 | 4 | 9 | 14 | 57 | 56 | 3.6 |  |  |
| **RSA212** | Male | 11 | Left | Left | 23 | 4 | 8 | 11 | 46 | 61 | 2.22 | 3.69 | 3.41 |
| **RSA215** | Female | 140 | Left | Left | 29 | 6 | 8 | 13 | 56 | 69 | 1 | 4.5 | 3.8 |
| **RSA216** | Male | 277 | Right | Left | 29 | 3 | 7 | 5 | 44 |  |  | 2.4 | 2.5 |
| **RSA217** | Male | 6 | Right | Right | 24 | 6 | 8 | 13 | 51 | 63 | 1.69 | 4.23 | 4.07 |
| **RSA220** | Female | 11 | Right | Right | 31 | 3 | 9 | 14 | 57 | 60 | 1.78 | 3.53 | 3.57 |
| **RSA221** | Male | 37 | Right | Right | 32 | 6 | 10 | 14 | 62 | 67 | 1.78 | 4.83 | 4.5 |
| **RSA223** | Male | 20 | Left | Left | 32 | 3 | 9 | 13 | 57 | 71 | 1.12 | 3.33 | 3.67 |
| **RSA225** | Female | 54 | Left | Right | 25 | 3 | 4 | 4 | 36 | 48 | 6.78 | 1.1 | 0.8 |
| **RSA226** | Female | 23 | Right | Right | 29 | 2 | 7 | 8 | 46 | 53 | 3.72 | 1.37 | 1.23 |
| **RSA227** | Male | 24 | Right | Left | 25 | 4 | 7 | 11 | 47 | 47 | 6.81 | 0.7 | 0.87 |
