## Supplemental Table S2 for "More Than Just Arm Movement: Finger-Worn Accelerometers Provide a Valid and Sensitive Alternative to Wrist-Worn Accelerometers for Measuring Real-World Upper-Limb Performance in Stroke Survivors"

Table S2: Summary of data completeness and adherence across study participants. Percentage of data retained and average daily wear time were calculated within days with complete 4-sensor data. Average daily wear time is reported as mean ± SD.

| **Subject** | **Total Days Recorded** | **Days with Complete**  **4-Sensor Data** | **Percentage of**  **Data Retained (%)** | **Average Daily Wear Time (h)** |
| --- | --- | --- | --- | --- |
| RSA201 | 8 | 8 | 85.12 | 6.73 ± 1.96 |
| RSA202 | 8 | 7 | 99.75 | 7.80 ± 1.49 |
| RSA203 | 8 | 5 | 95.09 | 11.80 ± 4.27 |
| RSA204 | 8 | 5 | 98.06 | 15.57 ± 8.56 |
| RSA205 | 9 | 7 | 99.47 | 9.62 ± 7.05 |
| RSA206 | 8 | 8 | 99.62 | 9.36 ± 2.78 |
| RSA207 | 8 | 8 | 99.44 | 9.49 ± 5.35 |
| RSA208 | 8 | 8 | 99.76 | 9.53 ± 2.56 |
| RSA209 | 8 | 5 | 98.17 | 7.69 ± 3.89 |
| RSA210 | 8 | 8 | 98.64 | 10.79 ± 4.10 |
| RSA212 | 8 | 8 | 99.78 | 7.00 ± 2.42 |
| RSA215 | 8 | 5 | 99.86 | 11.03 ± 3.07 |
| RSA216 | 8 | 7 | 99.52 | 6.43 ± 2.16 |
| RSA217 | 8 | 8 | 99.88 | 9.63 ± 2.71 |
| RSA220 | 9 | 9 | 80.98 | 9.66 ± 2.87 |
| RSA221 | 7 | 2 | 98.29 | 5.36 ± 4.09 |
| RSA223 | 7 | 4 | 92.92 | 9.37 ± 9.07 |
| RSA225 | 8 | 7 | 95.69 | 6.59 ± 2.37 |
| RSA226 | 7 | 5 | 94.57 | 9.34 ± 1.85 |
| RSA227 | 7 | 4 | 87.67 | 8.04 ± 1.18 |
| Total | 158 | 128 | 95.82 | 9.05 ± 4.52 |
