## Supplemental Table S3 for "More Than Just Arm Movement: Finger-Worn Accelerometers Provide a Valid and Sensitive Alternative to Wrist-Worn Accelerometers for Measuring Real-World Upper-Limb Performance in Stroke Survivors"

Table S3: Summary statistics of the wearable-based motor performance measures.

| **Wearable-Based Motor Performance Measures** | **Movement Time-Series** | **Descriptive Statistics** | | | | | | | |
| --- | --- | --- | --- | --- | --- | --- | --- | --- | --- |
|  |  | **Mean** | **Median** | **Min** | **Max** | **Range** | **Std** | **IQR** | **Skewness** |
| Average Intensity of Stroke-Affected Limb | Combined | 0.041 | 0.038 | 0.010 | 0.107 | 0.097 | 0.024 | 0.027 | 1.072 |
|  | Gross-Arm | 0.027 | 0.027 | 0.008 | 0.065 | 0.057 | 0.015 | 0.018 | 0.795 |
|  | Fine-Hand | 0.008 | 0.007 | 0.002 | 0.027 | 0.025 | 0.006 | 0.007 | 1.471 |
| Percentage Duration of Active Use of Stroke-Affected Limb | Combined | 0.213 | 0.210 | 0.041 | 0.473 | 0.433 | 0.124 | 0.189 | 0.449 |
|  | Gross-Arm | 0.160 | 0.147 | 0.021 | 0.333 | 0.312 | 0.089 | 0.131 | 0.286 |
|  | Fine-Hand | 0.077 | 0.060 | 0.014 | 0.255 | 0.241 | 0.064 | 0.081 | 1.302 |
| Use Ratio | Combined | -0.591 | -0.601 | -2.002 | 0.097 | 2.099 | 0.536 | 0.544 | -0.804 |
|  | Gross-Arm | -0.548 | -0.605 | -1.813 | 0.109 | 1.922 | 0.469 | 0.612 | -0.737 |
|  | Fine-Hand | -1.062 | -0.975 | -2.601 | 0.364 | 2.965 | 0.821 | 1.255 | -0.101 |
