## Supplemental Table S4 for "More Than Just Arm Movement: Finger-Worn Accelerometers Provide a Valid and Sensitive Alternative to Wrist-Worn Accelerometers for Measuring Real-World Upper-Limb Performance in Stroke Survivors"

Table S4: Mean and range of Spearman correlation coefficients between wearable-based motor performance measures and the FMA-UE across iterations of jackknife sensitivity analysis. Bold formatting indicates the strongest correlation coefficients among the combined, gross-arm, and fine-hand movement time-series.

| **Wearable-Based Motor**  **Performance Measures** | **Movement**  **Time-Series** | **FMA-UE** | | | | |
| --- | --- | --- | --- | --- | --- | --- |
|  |  | **Upper**  **Extremity** | **Coordination**  **& Speed** | **Wrist** | **Hand** | **Total** |
| Average Intensity  of Stroke-Affected Limb | Combined | 0.64 [0.59, 0.70] | 0.30 [0.19, 0.42] | 0.72 [0.68, 0.78] | 0.54 [0.49, 0.61] | 0.72 [0.68, 0.78] |
|  | Gross-Arm | 0.62 [0.56, 0.66] | 0.23 [0.11, 0.34] | 0.64 [0.59, 0.70] | 0.45 [0.39, 0.55] | 0.66 [0.60, 0.72] |
|  | Fine-Hand | 0.59 [0.54, 0.67] | 0.41 [0.33, 0.56] | 0.80 [0.76, 0.84] | 0.73 [0.70, 0.78] | **0.79 [0.77, 0.83]** |
| Percentage Duration of Active Use  of Stroke-Affected Limb | Combined | 0.59 [0.42, 0.68] | 0.37 [0.24, 0.50] | 0.72 [0.61, 0.81] | 0.54 [0.39, 0.66] | 0.72 [0.57, 0.81] |
|  | Gross-Arm | 0.63 [0.52, 0.69] | 0.26 [0.15, 0.42] | 0.68 [0.62, 0.74] | 0.49 [0.41, 0.57] | 0.68 [0.62, 0.75] |
|  | Fine-Hand | 0.56 [0.50, 0.68] | 0.45 [0.36, 0.56] | 0.78 [0.74, 0.83] | 0.71 [0.67, 0.76] | **0.78 [0.73, 0.82]** |
| Use Ratio | Combined | 0.54 [0.45, 0.65] | 0.33 [0.23, 0.44] | 0.71 [0.66, 0.82] | 0.59 [0.53, 0.69] | 0.69 [0.64, 0.79] |
|  | Gross-Arm | 0.51 [0.38, 0.59] | 0.22 [0.10, 0.32] | 0.60 [0.55, 0.70] | 0.46 [0.40, 0.59] | 0.59 [0.53, 0.68] |
|  | Fine-Hand | 0.61 [0.56, 0.67] | 0.37 [0.27, 0.48] | 0.79 [0.76, 0.83] | 0.72 [0.67, 0.78] | **0.78 [0.75, 0.81]** |
