## Supplemental Table S5 for "More Than Just Arm Movement: Finger-Worn Accelerometers Provide a Valid and Sensitive Alternative to Wrist-Worn Accelerometers for Measuring Real-World Upper-Limb Performance in Stroke Survivors"

| **Wearable-Based Motor**  **Performance Measures** | **Movement**  **Time-Series** | **WMFT** | | **MAL** | |
| --- | --- | --- | --- | --- | --- |
|  |  | **FAS** | **TIME** | **AOU** | **QOM** |
| Average Intensity  of Stroke-Affected Limb | Combined | **0.83 [0.80, 0.87]** | **-0.64 [-0.72, -0.58]** | 0.74 [0.70, 0.78] | **0.70 [0.67, 0.75]** |
|  | Gross-Arm | 0.77 [0.72, 0.86] | -0.58 [-0.67, -0.51] | 0.65 [0.59, 0.75] | 0.61 [0.56, 0.72] |
|  | Fine-Hand | **0.83 [0.80, 0.89]** | -0.59 [-0.68, -0.54] | **0.75 [0.73, 0.78]** | 0.69 [0.66, 0.75] |
| Percentage Duration of Active Use  of Stroke-Affected Limb | Combined | **0.84 [0.75, 0.92]** | **-0.67 [-0.74, -0.55]** | **0.76 [0.64, 0.82]** | **0.75 [0.66, 0.83]** |
|  | Gross-Arm | 0.81 [0.73, 0.86] | -0.64 [-0.72, -0.57] | 0.71 [0.63, 0.76] | 0.67 [0.60, 0.76] |
|  | Fine-Hand | **0.84 [0.78, 0.88]** | -0.63 [-0.67, -0.58] | **0.76 [0.73, 0.80]** | 0.72 [0.68, 0.76] |
| Use Ratio | Combined | **0.93 [0.89, 0.95]** | -0.73 [-0.77, -0.66] | 0.73 [0.68, 0.78] | 0.72 [0.65, 0.76] |
|  | Gross-Arm | 0.89 [0.87, 0.92] | **-0.75 [-0.82, -0.71]** | **0.77 [0.74, 0.83]** | 0.70 [0.66, 0.76] |
|  | Fine-Hand | 0.90 [0.87, 0.95] | -0.68 [-0.76, -0.62] | 0.75 [0.71, 0.81] | **0.75 [0.69, 0.80]** |

Table S5: Mean and range of Spearman correlation coefficients between wearable-based motor performance measures and the WMFT and MAL across iterations of jackknife sensitivity analysis. Bold formatting indicates the strongest correlation coefficients among the combined, gross-arm, and fine-hand movement time-series. Since shorter task completion times reflect better upper-limb performance,^37^ negative correlations were found between wearable measures and WMFT-TIME.
