## Supplementary material for "More Than Just Arm Movement: Finger-Worn Accelerometers Provide a Valid and Sensitive Alternative to Wrist-Worn Accelerometers for Measuring Real-World Upper-Limb Performance in Stroke Survivors": Jackknife Analysis

Jackknife Sensitivity Analysis

A jackknife sensitivity analysis (i.e., leave-one-out resampling) was performed to assess the robustness of statistical estimates to sampling variability. Specifically, one participant was systematically excluded at a time, and the optimal acceleration thresholds *α*, Spearman correlation coefficients, and ICC(2,1) values were then recalculated across the remaining 19 participants over 20 iterations. Mean and range of correlation and ICC values were reported across all iterations.

Results

The mean correlation and ICC values across iterations of the jackknife sensitivity analysis were consistent with the results from the full-sample analysis, with small variability (Supplemental Table S4, Supplemental Table S5, and Supplemental Figure S4). These findings support the robustness of the reported correlations and ICC values to sampling variability.
